# Competing Risk Model in Dental Cost-Effective Research

**DOI:** 10.1101/2025.01.30.25320359

**Authors:** Anand Seth, Muhammad H.A. Saleh, Priyanshi Shah, Arinjita Bhattacharyya, Hamzeh Almashni, Guiseppe Troiano, Himabindu Dukka, Shesh N. Rai

## Abstract

Over the years, few attempts have been made to use survival analysis techniques in dental research. Moreover, the use of survival analysis was restricted to a simple method of drawing a survival curve using the Kaplan-Meier technique, but later prognostic factors were included utilizing the Cox Proportional Hazard (CPH) model. More recently, the CPH model has been incorporated with a frailty parameter to account for the correlation of observations within a subject. A recent publication has utilized a subdistribution hazard model with time-dependent covariates. In actual dental practice, a tooth failure could be due to a variety of reasons other than the cause of failure of interest. To have a realistic estimate of the risk of failure for cause i, one should also include the risk of failure due to causes j, j=1(1) n, which could occur prior to cause i. This competing risk scenario has been attempted in the last ten to fifteen years or so to estimate the probability of failure in the presence of competing event(s). Precise estimate of survival (failure) probabilities using competing risk model has implications in the dental cost-effectiveness and cost-utility studies.

## Introduction

Over the years, few attempts have been made to use survival analysis techniques in dental research. Between 1996 to 2007, an increase in survival analysis methods was reported [8]. In a study of fifty-three compomer restorations, the probability that a restoration would be clinically acceptable was determined to be 89% using the Kaplan-Meier (KM) method [9]. Zimmer et al. showed Cerec (Chairside Economical Restoration of Esthetic Ceramic) restorations were comparable to cast gold restorations using KM method [10]. KM survival curves were used to estimate the survival of first permanent molars caries-free, whether sealed or non-sealed in two centers in Finland [10], [11]. A prospective study on inlay-retained fixed dental prosthesis, over an average period of 70 months, survival of 57% after 5 years and 38% after 8 years using KM method was shown [12], [13]. Opdam et al. 2011 recommended the KM method as a method of choice for calculating the longevity of a group of dental restorations [12]. To account for the effect of covariates, a semiparametric approach called Cox proportional hazard (CPH) was used [13]– [15]. Kim et al. 2013 compared longevity of restoration of amalgam, composite resin, and glass ionomer using the KM and CPH methods [16]. More recently, a randomized control study was conducted in 65 years or older adults with partial dentate comparing two groups utilizing the CPH model [17]. Survival probabilities were computed under primary crown retained removable partial studied in a group of 653 patients who received 953 post and cores using the KM and CPH methods [18]. The first attempt to apply the CPH model with frailty (correlation of observations within a same patient) appears to be by Pinto et al. (2014) [3]. A recent publication related to 1275 direct composite restorations in 185 patients applied CPH model with a frailty parameter to account for restorations within the same patient [5]. In a similar study of 9184 restorations among 1551 unique patients, a CPH model with patient-level random effect (frailty) was fitted [4].

In 2012, Pajewski et al. applied the competing risk (CR) model to predict the probabilities of returning to the Emergency Department following a non-traumatic dental condition versus obtaining follow-up care from a dentist [19]. There has been recent interest in applying CR models in dental research [20]. Periodontal disease and the incident of coronary heart disease is explored using a CR model in a large cohort of 6300 patients [21]. Zhang et al. recently explored periodontitis and risk of diabetes correlation using a CR model [22]. A 2023 publication has utilized a subdistribution hazard model with time-dependent covariates [6]. A quantity commonly used in survival analysis is hazard rate. Hazard rate can be thought of as the probability of losing a tooth at time t if that tooth has survived just prior to time *t*. A precise estimate of hazard rate is helpful in correctly estimating the cost of providing future dental care, especially periodontal care, over a long follow-up period. In medical and dental studies, often, there are multiple causes of an event. During supporting periodontal therapy (SPT), a patient may experience a periodontal loss, but it is also possible that a patient may experience a non-periodontal loss before a periodontal loss. Such a situation is often referred to as a “competing risk” scenario. In dental research literature, often, this competing risk scenario is ignored while computing hazard rates at different follow-up times. This often results in an overestimation of the probability of losing a tooth, which in turn will affect the estimated cost over a long course of periodontal treatment. Note that estimating the probability of losing a tooth due to periodontitis correctly is critical in developing a realistic cost estimate since other causes of tooth losses over time could and do influence this probability and, hence, the cost. The aim of the present paper is to estimate hazard rate λ (t) and an associated quantity Cumulative Incidence Function (CIF) under the competing risk scenario. CIF represents cumulative probability of tooth loss while accounting for losses other than periodontal loss. The hazard rate λ (t) can be estimated by non-parametric, semi-parametric, and parametric biostatistical methods. Therefore, the aim of this study was to provide recommendations for computing λ (t) and CIF under different scenarios while utilizing retrospective data to compute hazard rates associated with different prognosticators for future cost-effective studies.

## Material and Methods

A patient may go from a healthy state to an unhealthy state (−1) to Miller and McEntire (MM) class II or III (state 0) and then to a state where he/she loses a tooth due to periodontal loss or due to some other reasons (state 1 or state 2). Such a “transitional” model is illustrated using the concept of state space (transitional probabilities). The state space framework allows great flexibility in modeling the outcome.

In Figure 1, a transition of different states is presented to illustrate the importance of different states and of incorporating competing risks in the determination of the probability of losing a tooth due to periodontal loss or other types of losses. One could further subdivide the other losses into more categories.

**Figure 1:**
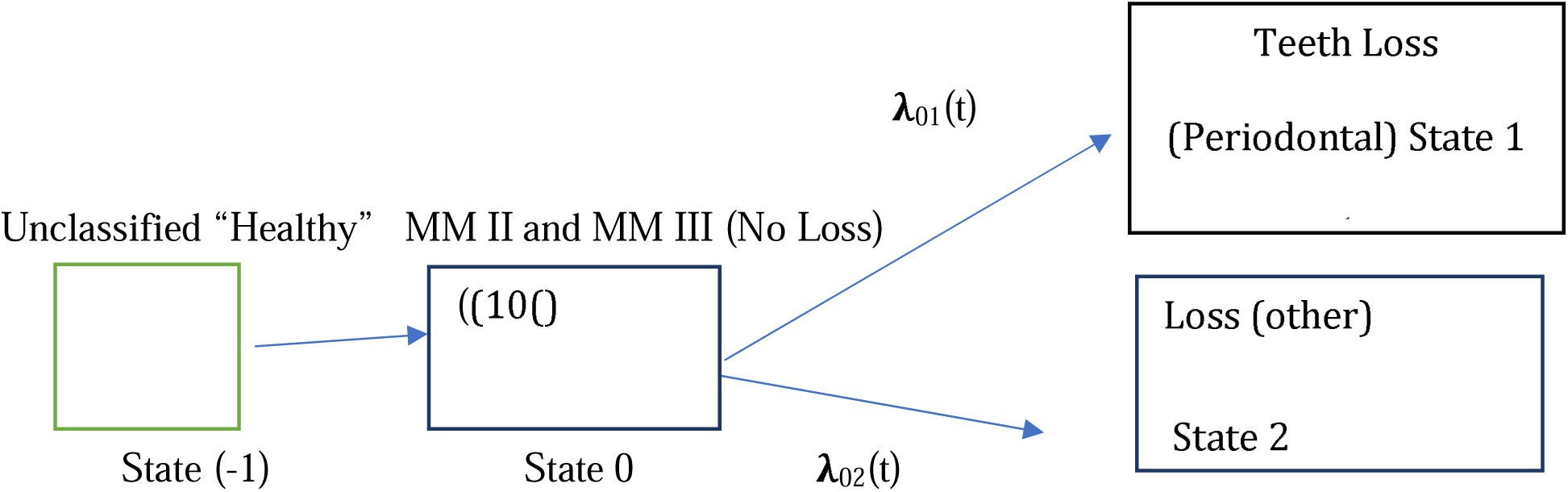
State-space Model of Periodontal Tooth Loss in Presence of Competing Risk

In the initial state space (−1), a patient is not classified as MM II to MM III. In the state space (0), a patient has been classified as MM Class II or III but has not experienced any tooth loss yet. Next, a patient moves from state (0) to either state (1) or state (2). λ_01_(t) represents a movement from state (0) to state (1) with an intensity λ_01_(t). This intensity represents a probability of going from a healthy state to a state where a patient experiences periodontal loss. Similarly, λ_02_(t) represents a patient going from state (0) to a state where they experience loss other than periodontal loss. These two states represent competing causes a given patient may experience. Since a competing cause is always present in the actual practice, it is necessary to take into consideration the competing cause(s) while we evaluate the survival probabilities or cause-specific (CS) cumulative incidence over time.

A common method of estimating of λ (t), is the non-parametric method of Kaplan and Meier [1]. Often, prognostic variables like age, sex, and number of maintenance visits etc., influence the hazard rate, survival, or mortality rate (See equation 1). To accommodate predictors, the Cox Proportional Hazard (CPH) model with covariates is used [16]. In the CPH model, the relative effect of a covariate can be assessed on the hazard function. The CPH model is often referred to as semiparametric model since λ(t), the baseline hazard is estimated non-parametrically and the other portion 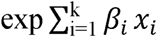 is estimated by parametric methods. Hazard function denotes an instantaneous rate of occurrence of an event i.e., periodontal loss at time t.

The CPH model is expressed as

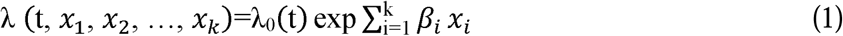

where λ_0_ (t) denotes baseline hazard at time t and *x*_1_, *x*_2_, …, *x*_k_ are independent prognostic variables or covariates as indicated above.

Note λ_0_ (t) is a non-parametric portion in (1) since no parametric distribution is imposed on λ_0_ (t). In the original formulation of CPH model, no parametric distribution was imposed on λ_0_ (t), but later parametric and some other types of structures were imposed on λ_0_ (t). Further, in our investigation, we will model λ_0_ (t) by the parametric models like exponential, Weibull, log-logistic, and log-normal to further improve “precision” in estimating hazard function.

In the CPH model, the key assumptions are that the effect of a covariate is same at all time points and that the ratio of hazards for any two subjects i and j is independent of time, often referred to as an assumption of proportionality.

An extension of the CPH model is available where the effect of a covariate can change over time. For details, refer to Austin et al. [23] and Zhang et al. [24]. It should be noted that when the hypothesis of proportionality is rejected, then one can use time-dependent covariates to assess hazard λ (t).

In general, the CPH model overestimates hazard function as compared to a competing risk model, which takes competing events into consideration while estimating λ_0_ (t). This model is called the Fine and Grey (FG) [7] model. Note that a patient may experience a non-periodontal tooth loss before he/she experiences a periodontal tooth loss. Accounting for such a loss does provide a more precise estimate of hazard rate for a periodontal loss. In the CPH model, competing events are treated as censored observations, but in the FG model, that is not the case. The models described here are referred to as non-parametric and semiparametric models. One can also use parametric models or a mixture of parametric models to model baseline hazard rate λ_0_ (t), which will be discussed in later sections.

### Competing Events

A competing risk (CR) is an event whose occurrence precludes the occurrence of the primary event of interest [7]. Why competing risks should be taken into consideration while estimating hazard rate λ(t) can be visualized using a State-Space Model of periodontal tooth loss in the presence of competing risk (Figure 1). A subject can go from a healthy state to a diseased state, which could be a periodontal loss or a non-periodontal loss. A periodontal tooth is extracted only after examining patient treatment notes, final X-ray, and a patient chart. A non-periodontal loss could be due to several reasons such as physical wear, low grade trauma, diabetes, smoking, cardiovascular disease, chronic obstructive pulmonary disease, and arthritis. The third state could be no loss within the observed time. The risk to a patient to go into different states can be expressed in terms of hazard (instantaneous risk of an event) or intensities, as shown in figure 1. These intensities λ_ij_(t) will indicate a person’s risk of going from state i to j. These intensities λ_ij_(t) are modeled either non-parametrically or parametrically as indicated above.

The hazard or intensity can be modeled semi parametrically using (1) and parametrically by using the following models:

Note that λ (t) is the ratio of the density f(t) to the survivor function S(t), λ(t) = f(t)/S(t).

In other words, the hazard function is related to the survivor function by a simple relationship.

The association of prognostic variables and outcome of interest can result in overestimation of Cumulative Incidence Function (CIF) if competing risks are present [7]. Fine and Grey [7] method allows us to estimate the subdistribution (SD) hazards for competing risk models, which denotes the instantaneous risk of failure from the k^th^ event type in subjects who have not experienced an event of type k. This method estimates the effect of covariates on the cumulative incidence function of the event of interest while accounting for failure due to other causes, and hence not overestimating, in general, the risk of loss or death of an event of interest.

Often, in dental studies, there is an event of interest like periodontal loss, but a patient may experience non-periodontal loss prior to a periodontal loss due to a variety of reasons. Non-periodontal loss, in this case, is considered a competing event. FG model correctly estimates the probability of a periodontal loss while accounting for a possible non-periodontal loss within the same cohort of patients using the CR model. A quantity called CIF summarizes the cumulative failure rate over time due to a particular cause. CIF estimates the marginal probability of each competing event. Since the computation of marginal probability does not assume the independence of competing events, it has relevance in dental cost-effectiveness and/or cost-utility analyses where risk probability is used to assess treatment utility. For our research question, the effect of covariates (age, sex, follow-up time, condition of tooth in terms of Miller and McEntire classification, and number of maintenance visits) is assessed on CIF.

### Description of Sample Data

The data for the current study were retrospectively collected between July 2019 and July 2021 from electronic and physical charts of patients receiving periodontal treatment at the University of Michigan School of Dentistry, Ann Arbor, Michigan, USA.

A total of 270 patients with advanced periodontitis were screened, of which 148 patients fulfilled the inclusion criteria. Excluded patients had incomplete data (charts, follow-up, or the cause of tooth loss) was not identified. Eighty females, 68 males, making a total of 3787 teeth, were included in the original analysis [25] with a mean age of 46.49 ± 11.53 years. At baseline, 1886 (49.6%) maxillary and 1901 (50.2%) mandibular teeth were present. For the current dataset, 373 (40.2%) maxillary and 141 (15.2%) mandibular teeth were present.

Out of the 3787 teeth, 937 teeth (24.7%) were either Miller/ [Periodontal risk score] (PRS) Class 2 or 3. The number of anterior teeth was 415 (44.2%), and posterior teeth were 522 (55.7%). MM Class 2 applied to 807 teeth (86.1%) and MM Class 3 applied to 130 teeth (13.9%). Over an average follow-up of 297.7 ± 91.3 months (24.7 ± 7.6 years), 212 teeth were lost (22.6%), of which 114 (12.2%) were lost due to periodontitis, and 98 (10.4%) were lost for reasons other than periodontitis. Data on 926 teeth was analyzed since nine teeth had missing data on the variable RE6 (SRP 1-3 teeth) and age, so these observations were excluded from all the subsequent analyses. In addition, one 17year old with two teeth with missing RE6 value was excluded from the analysis. Of all the excluded teeth, three had “other loss’ status.

## Results

Table 2 below provides characteristics of a Retrospective Sample

**Table 1:** Common Parametric Models to Characterize Hazard at Time t.

| Distributions | Density f(t) | Hazard $\lambda(t)$ | Survival S(t) |
| --- | --- | --- | --- |
| Exponential | $\alpha e^{-\alpha t}$ | $\alpha$ | $e^{-\alpha t}$ |
| Weibull | $\alpha \beta t^{\beta-1} e^{-\alpha t^\beta}$ | $\alpha \beta t^{\beta-1}$ | $e^{-\alpha t^\beta}$ |
| Log-Logistic | $\frac{\alpha \beta t^{\beta-1}}{[1 + \alpha t^\beta]^2}$ | $\frac{\alpha \beta t^{\beta-1}}{1 + \alpha t^\beta}$ | $\frac{1}{1 + \alpha t^\beta}$ |
| Log-normal | $\Phi\left\{\frac{\ln\{t\} - \alpha}{\beta}\right\}$ | $\frac{f(t)}{S(t)}$ | $1 - \Phi\left\{\frac{\ln\{t\} - \alpha}{\beta}\right\}$ |

Note: Parametrization of density functions shown above is commonly used in medical research

**Table 2:** Characteristics of Teeth.

|  | Total Teeth<br>(%) | No Loss | Other<br>Loss | Perio<br>Loss | P<br>values<sup>+</sup> |
| --- | --- | --- | --- | --- | --- |
| Age of Teeth (yrs.) | N=926 | N=717 | N=95 | N=114 |  |
| < 25 (1) | 17 (1.8) | 10 (58.8) | 0 (0.0) | 7 (41.2) | 0.001 |
| 25-49 (2) | 543 (58.6) | 432<br>(79.6) | 50 (9.2) | 61 (11.2) |  |
| 50-69 (3) | 333 (36.0) | 245<br>(73.6) | 43 (12.9) | 45 (13.5) |  |
| ≥ 70 (4) | 33 (3.6) | 30 (90.9) | 2 (6.1) | 1 (3.0) |  |
| N | 926 | 717<br>(77.4) | 95 (10.3) | 114 (12.3) |  |
| Mean ± SD (Yrs.) | 46.3±12.0 | 46.3±12.1 | 48.6±11.4 | 44.3±11.9 |  |
| Min, Max (Yrs.) | 24, 76 | 24, 76 | 27, 76 | 24, 71 |  |
| Mean ± SD (# of Visits –<br>Maintenance) | 58.9±16.3 | 33.2±19.7 | 32.8±20.6 |  |  |
| Min, Max | (16, 101) | (1, 79) | (2, 83) |  |  |
| Sex |  |  |  |  |  |
| Male (1) | 499 (53.9) | 387<br>(77.6) | 46 (9.2) | 66 (13.2) | 0.382 |
| Female (0) | 427 (46.1) | 330<br>(77.3) | 49 (11.4) | 48 (11.2) |  |
| MM Class |  |  |  |  |  |
| 2 (good) | 796 (86.0) | 640<br>(80.4) | 77 (9.7) | 79 (9.9) | <0.0001<sup>+</sup> |
| 3 (guarded) | 130 (14.0) | 77 (59.2) | 18 (13.9)) | 35 (26.9) |  |
| Status of Tooth |  |  |  |  |  |
| Upper Molar | 371 (40.1) | 273<br>(73.6) | 45 (12.3) | 53 (14.3) | 0.005 |
| Lower Molar | 141 (15.2) | 100<br>(70.9) | 18 (12.8) | 23 (16.3) |  |
| Non-Molar | 414 (44.7) | 344<br>(83.1) | 32 (7.7) | 38 (9.2) |  |
| Total | 926* |  |  |  |  |

FUP (years) Mean =24.5; Median= 24, Min =0.0833, Maximum=4.5 <sup>+</sup>7P<0.0001 for Perio vs. non-periodontal., + \* Nine teeth were excluded due to missing information on a variable RE6, in addition, one subject, a 17-year-old with two teeth was excluded from the analysis due to missing information on variable RE6; <sup>+</sup> Chi-Square.

In this retrospective sample, age and type of loss showed high significance (p=0.001), and loss of a tooth was highly related to the Miller class (p=0.0001). The incidence of periodontal disease was more than twice (26.9%) in MM class III vs. MM class II (9.9%). Also, the loss of a tooth depended on whether the tooth was an upper molar, lower molar or non-molar (p=0.005). Overall, the relationship between sex and loss of tooth was not significant (p=0.382).

Table 3 gives cumulative hazard rate (cumulative mortality) by Kaplan and Meier and Nelson-Aalen (NA) methods for 30 years follow up time.

**Table 3:**
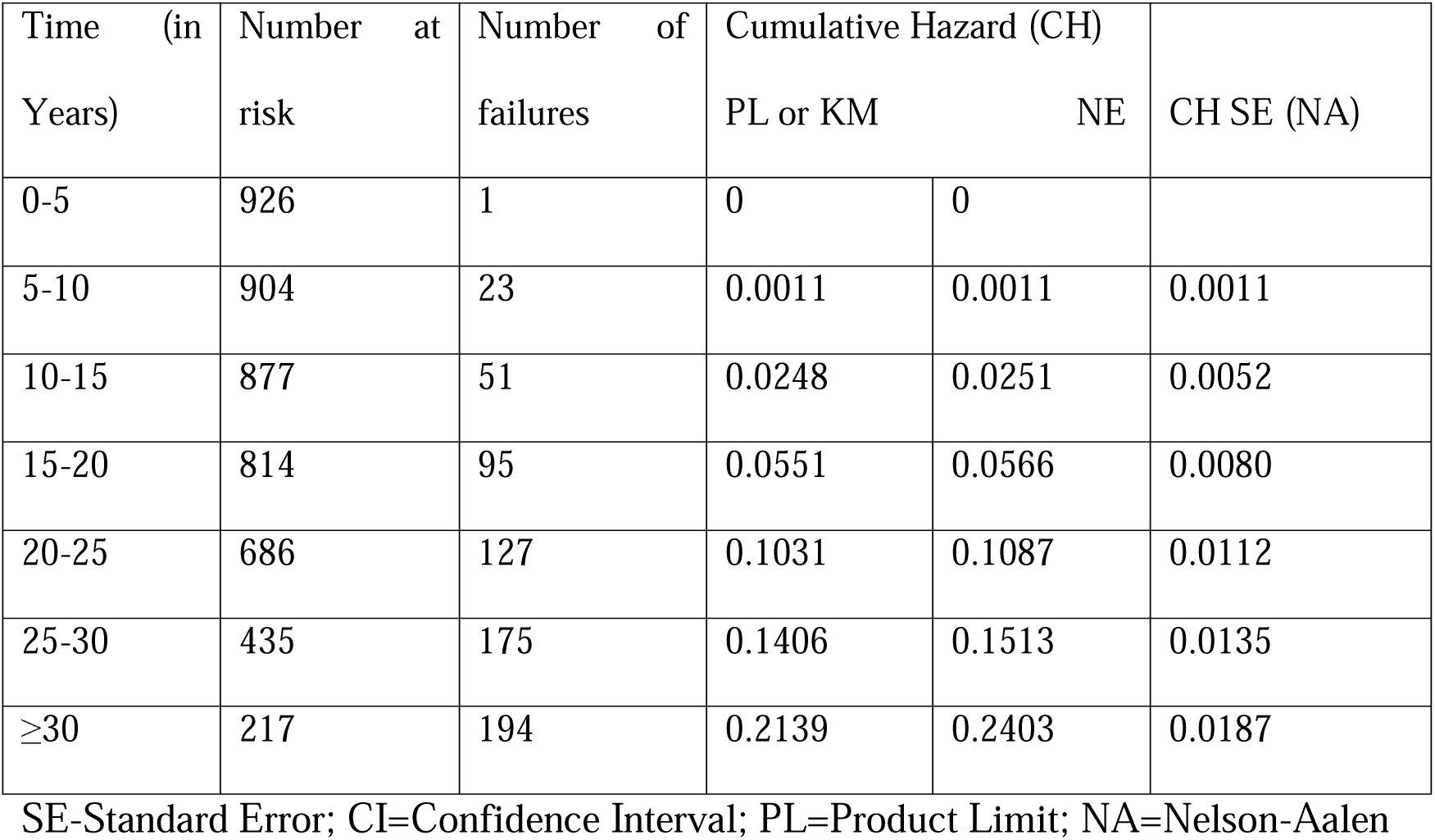
Overall Survival Probability of teeth using KM Method with a 30 year follow up by age of participants between 24-80 years.

**Table 4:**
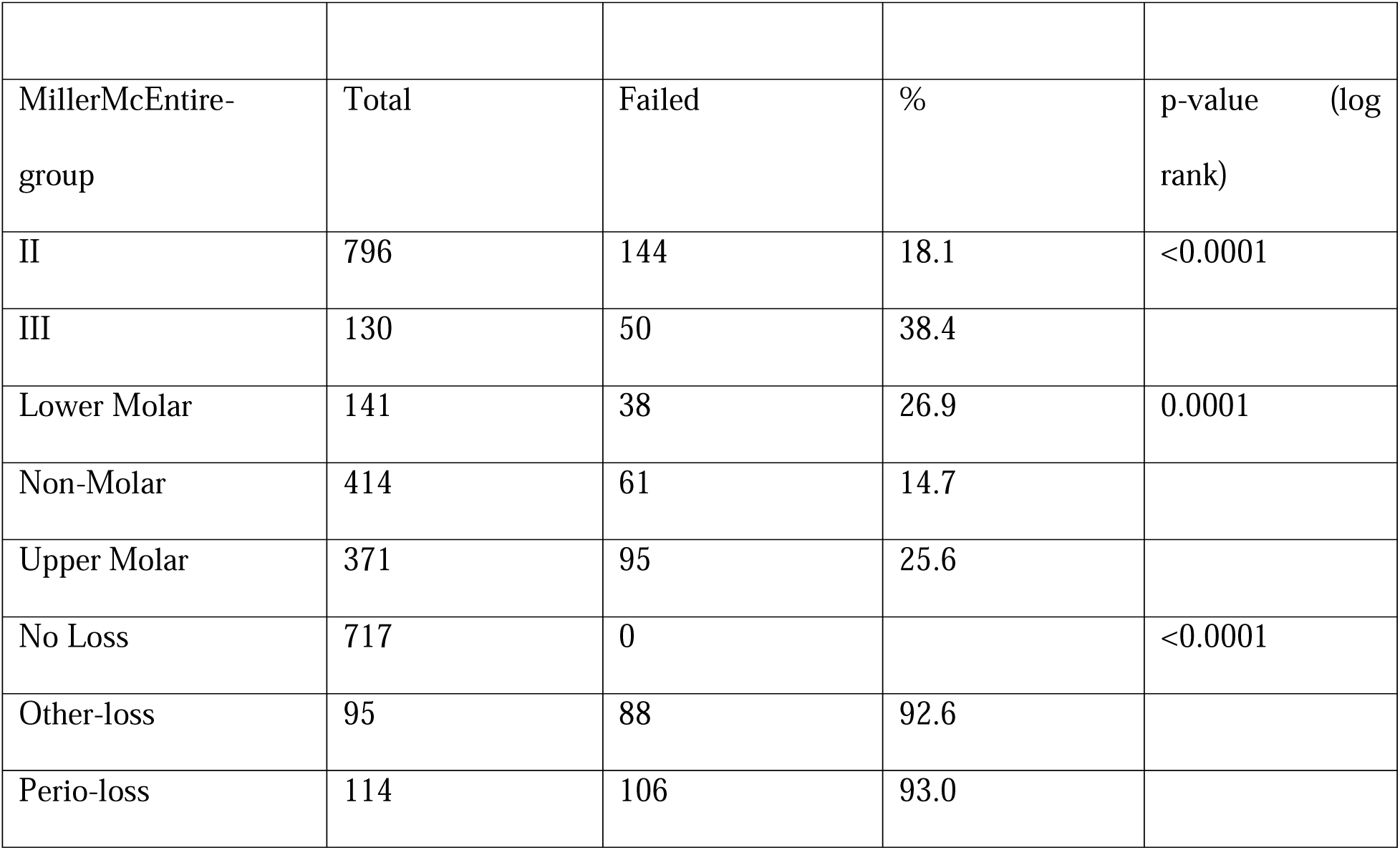
Failure of Teeth by MM Class, Type of Teeth, and Status over a Follow-up Period of 30 Years using the Kaplan-Meier Model.

From Figure 2, we see that the survival probability of teeth decreases with time much more for MM class III than for MM class II.

**Figure 2:**
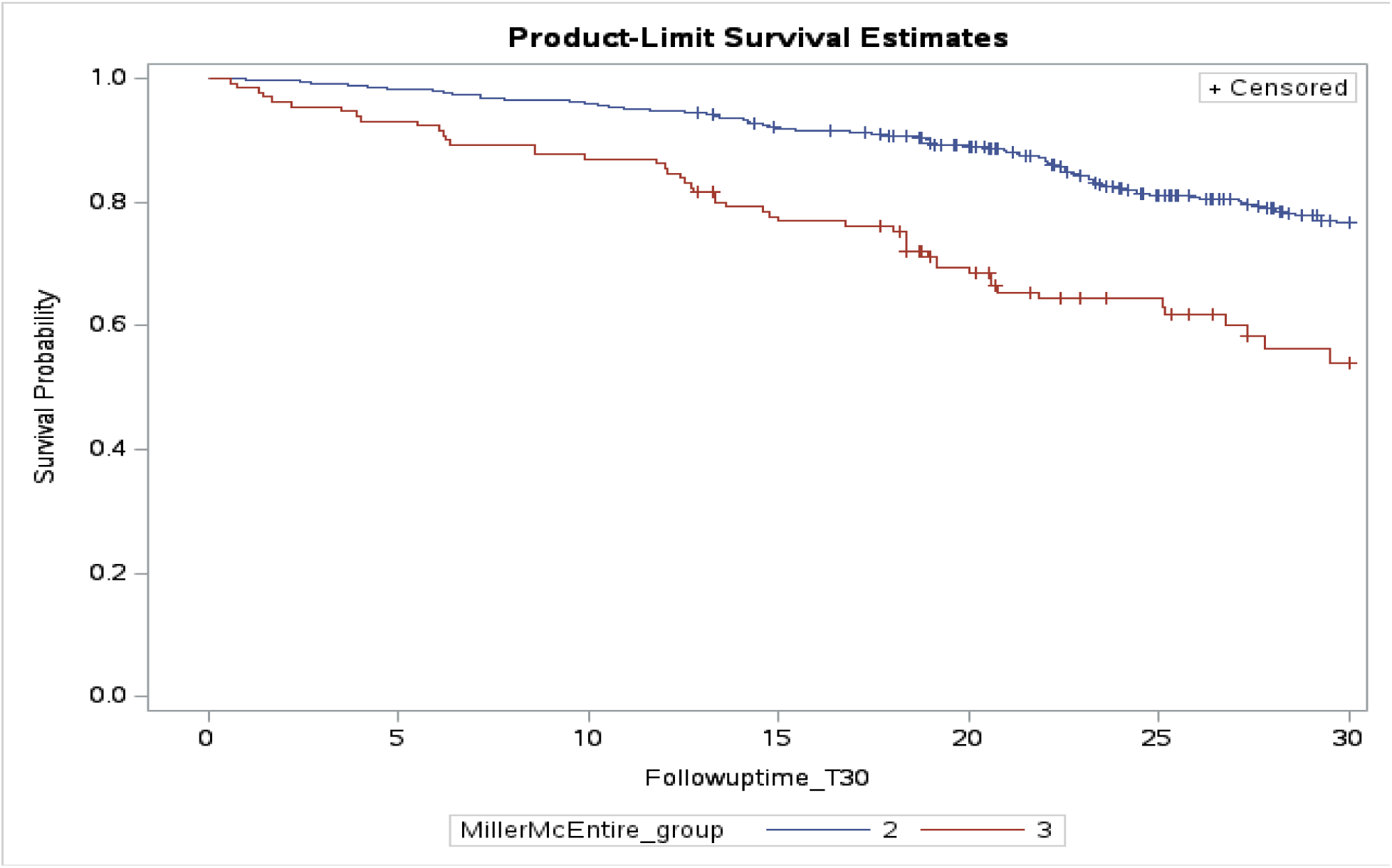
KM Survival Probability of Periodontal Loss by MM Class Over a Follow-Up Period of 30 Years

In Figure 3, non-molar has a higher probability of survival than lower and upper molar, both of which tend to follow a similar path.

**Figure 3:**
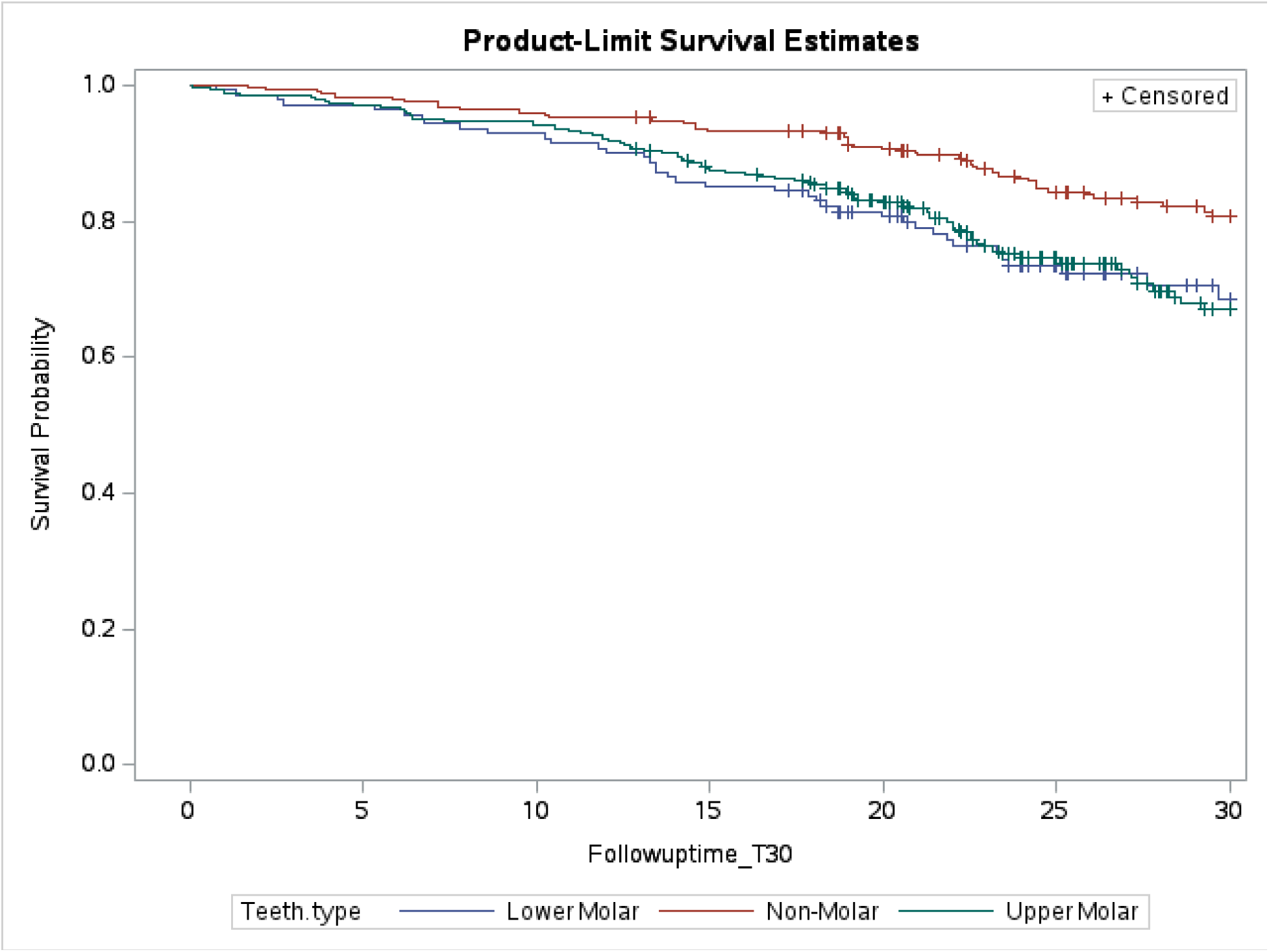
KM Survival Probability of Periodontal Loss by Teeth Type Over a Follow-Up Period of 30 Years

**Figure 4:**
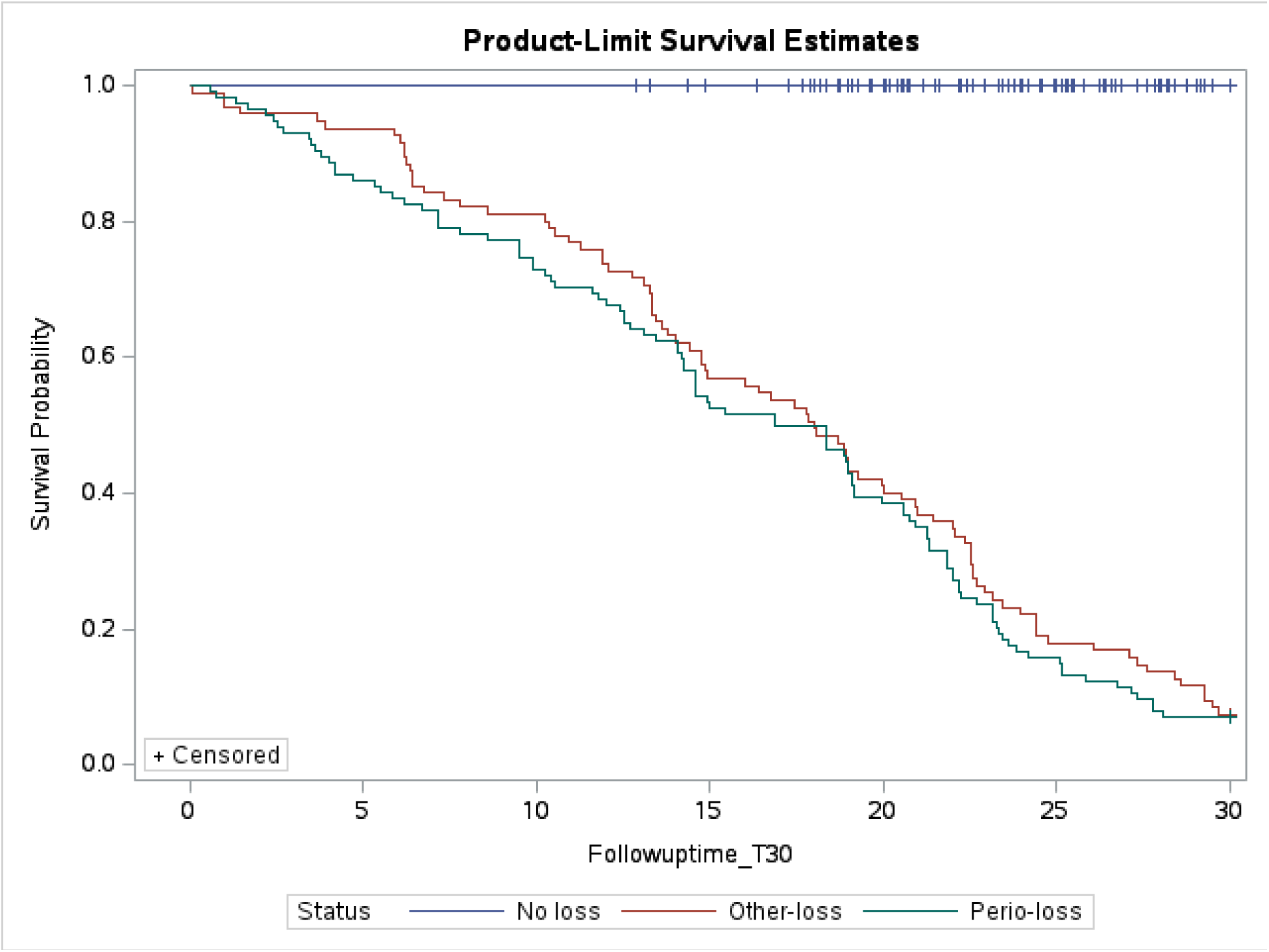
KM Survival Probability of Periodontal Loss by Status Over a Follow-Up Period of 30 Years

From the above figure, it is clear that periodontal-loss survival probability is lower than that of other-loss.

Results of MM class shows much higher percentage of failure 38.4% vs. 18.1% (p<0.0001). Also, teeth type (p=0.0001) and the three categories no-loss, other-loss and periodontal-loss were statistically significantly different (p<0.0001).

Table 5 provides the number at risk by type of teeth in a 5-year interval over a period of 30 years.

**Table 5:** Cox proportional hazard model with Follow up time of 30 years and age between 24-80 years.

| Teeth Type | Lower Molar (%) | Upper Molar (%) | Non-molar (%) | Total (%) |
| --- | --- | --- | --- | --- |
| Number at Risk<br>(in Years) |  |  |  |  |
| 0 | 141 (100) | 371 (100) | 414 (100) | 926 (100) |
| 5 | 137 (97.2) | 360 (97.0) | 407 (98.1) | 904 (97.6) |
| 10 | 131 (92.9) | 349 (94.1) | 397 (95.9) | 877 (94.7) |
| 15 | 120 (85.1) | 318 (85.7) | 374 (90.3) | 812 (87.7) |
| 20 | 97 (68.8) | 269 (72.5) | 307 (74.2) | 673 (72.7) |
| 25 | 65 (46.1) | 151 (40.7) | 175 (42.3) | 391 (42.2) |
| 30 | 35 (24.8) | 82 (22.1) | 110 (26.6) | 227 (24.5) |

Since in cost-effective/cost-utility studies, time horizon is normally taken to be 30 or 36 years, hence we present the result of this analysis to a 30 year follow up time. Table 6 shows the results based on status (periodontal vs. non-periodontal loss) regressed with covariates through the CPH model and competing risk model (cause-specific and subdistribution), with a follow up period of 30 years, respectively. Age (p_sd_=0.0006) was significant. In addition, maintenance (p_sd_ <0.0001) and MM class (p_sd_ =0.004) both are significant by the PH, cause-specific and subdistribution models. Risk of losing a tooth is approximately 2 ½ times greater if in MM class III as compared to in MM class II (p_sd_=0.004). Note that HR is 3.36 for MM class III by the cause-specific model while 2.28 with the subdistribution CR model, a considerable overestimate of risk (3.36 vs. 2.28). Though not statistically significant at the 0.05 level, risk of losing lower molar is consistently higher by all the three models with a 41% higher chance of losing a lower molar tooth compared to non-molar tooth. Proper maintenance (number of visits) of teeth does reduce the chance of losing a tooth (p<0.001).

**Table 6:**
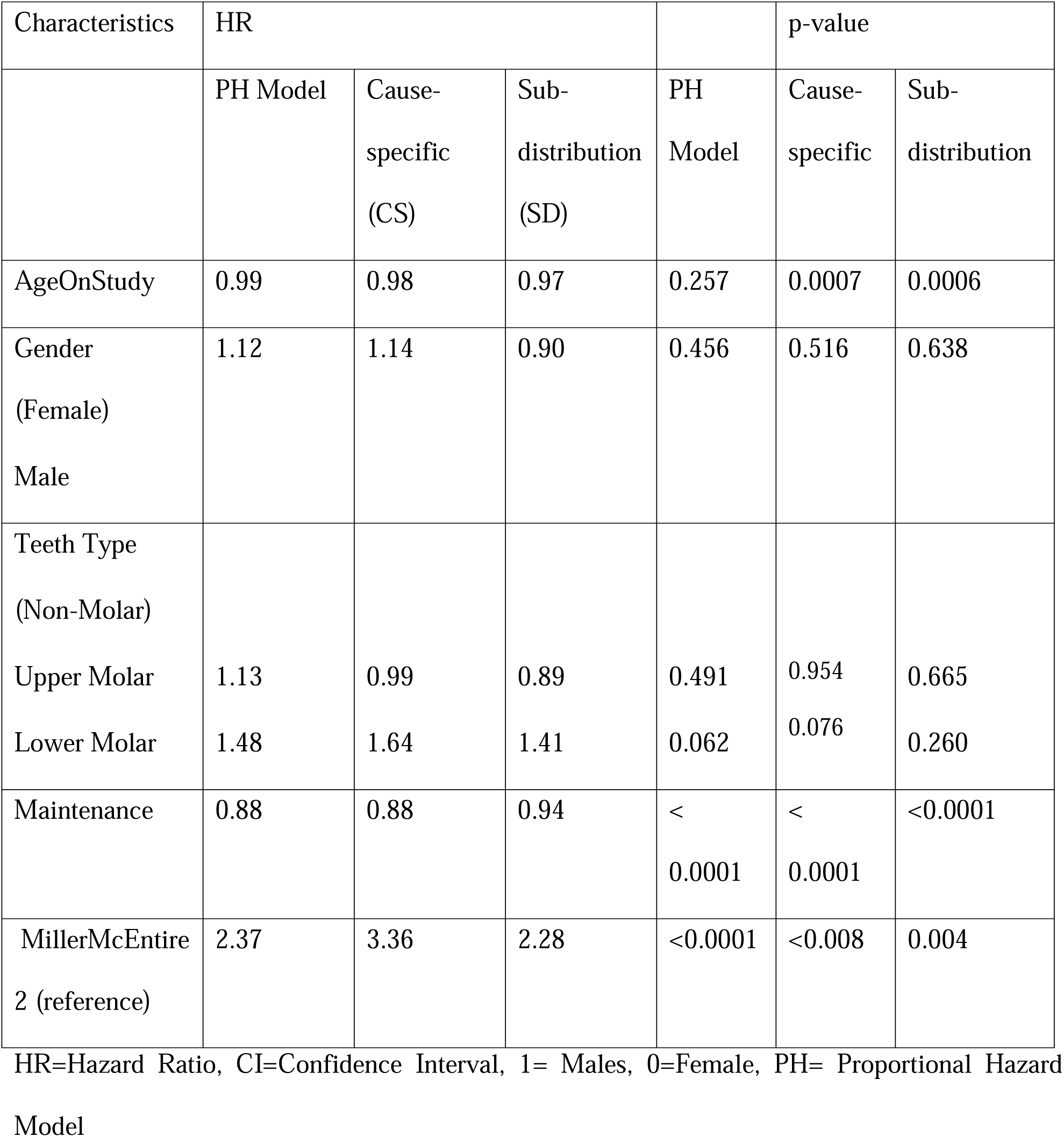
Cox Proportional hazard and Fine-Grey model with Follow up time of 30 years and age between 24-80 years.

Figure 5 gives CIF of periodontal loss by MM class. Clearly, cumulative hazard of losing a tooth of class III is much higher than class II.

**Figure 5:**
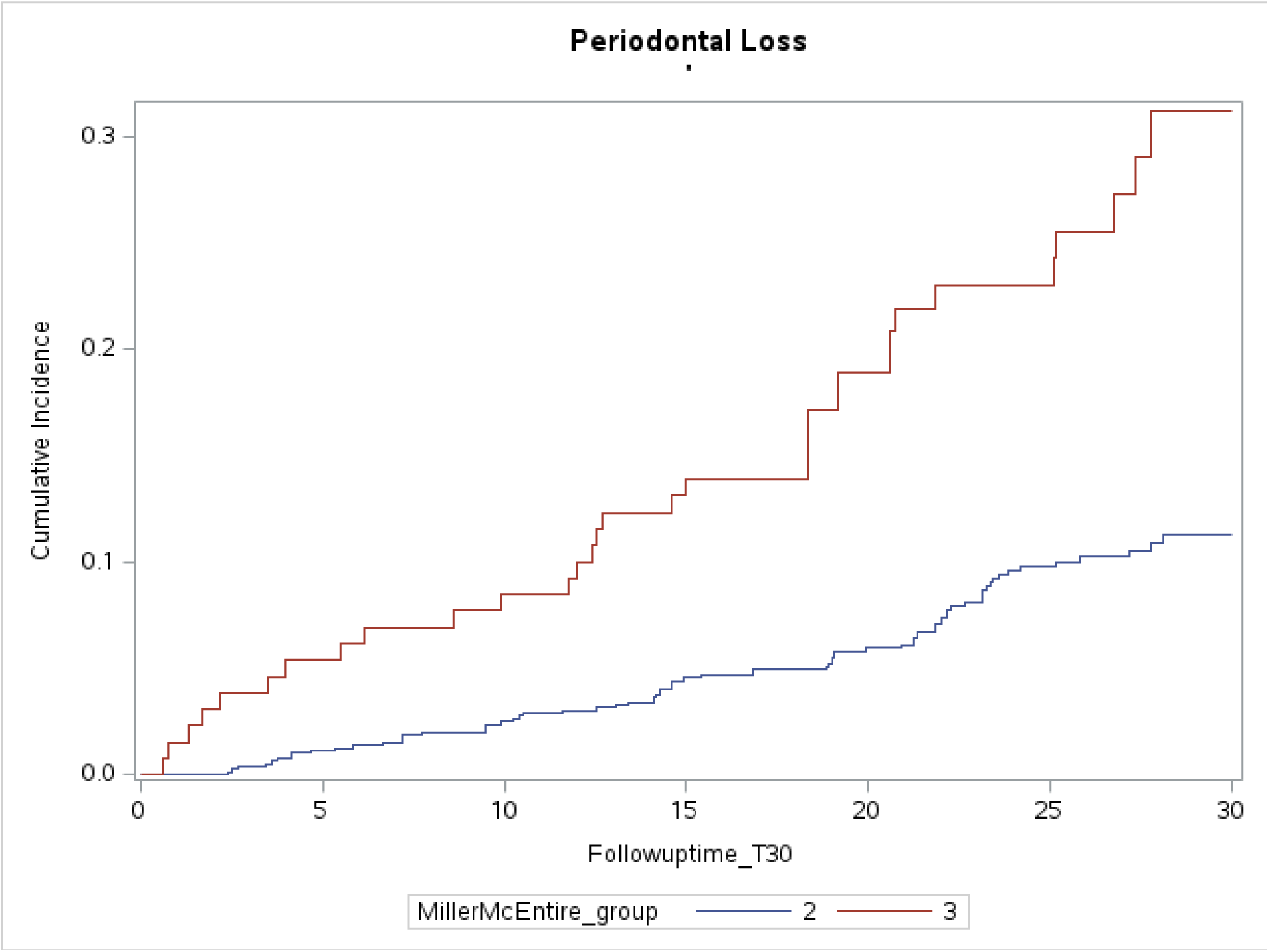
Cumulative Incidence Function (CIF) of Periodontal Loss over a 30 Year Follow Up Period by Periodontal Risk Score

**Figure 6:**
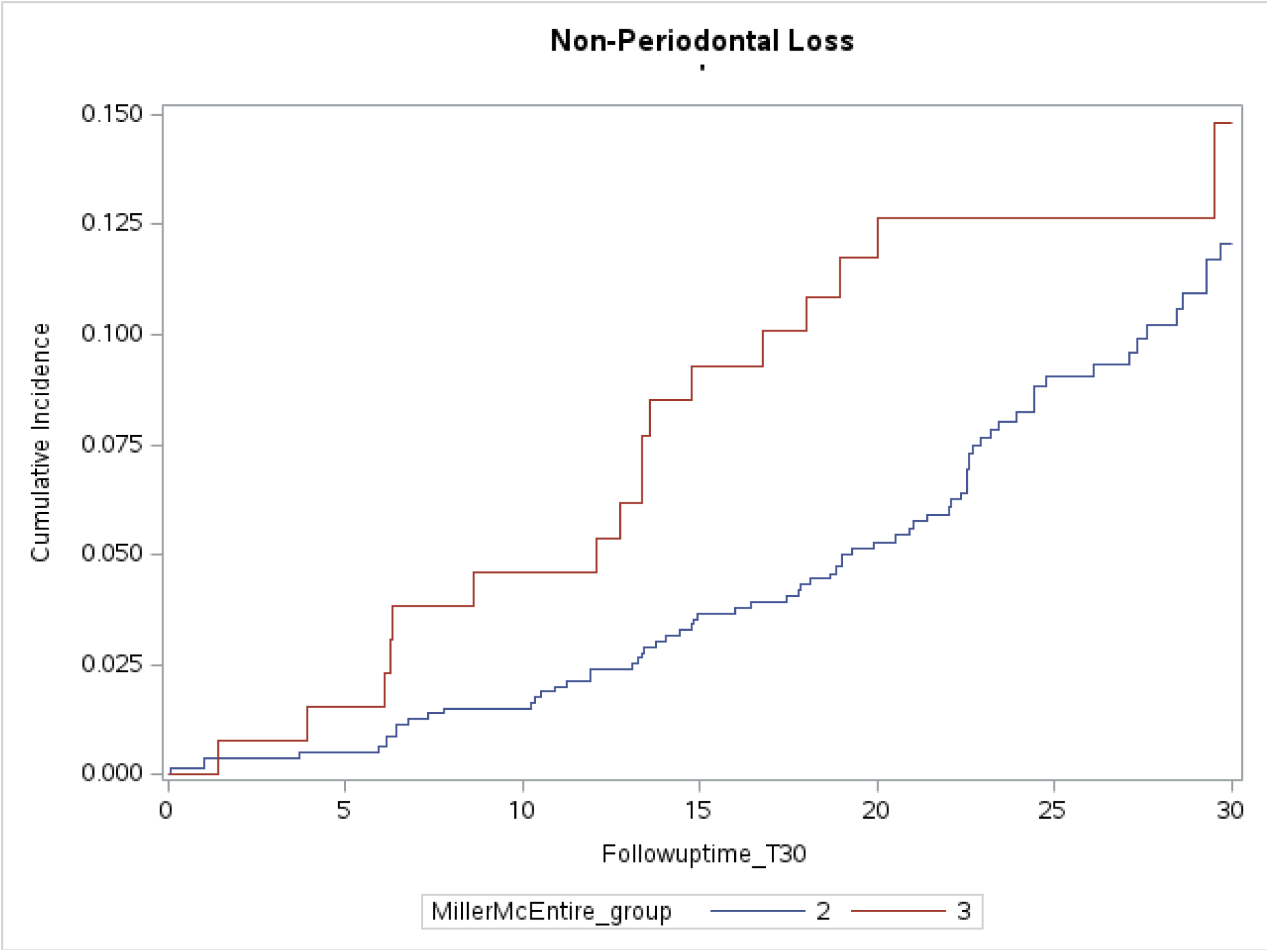
Cumulative Incidence Function (CIF) of Non-Periodontal Loss over a 30 Year Follow Up Period by Periodontal Risk Score

For non-periodontal loss, again CIF for MM class III is much higher throughout than MM class II.

Figure 7 gives overall CIF of periodontal loss over a 30 year follow up.

**Figure 7:**
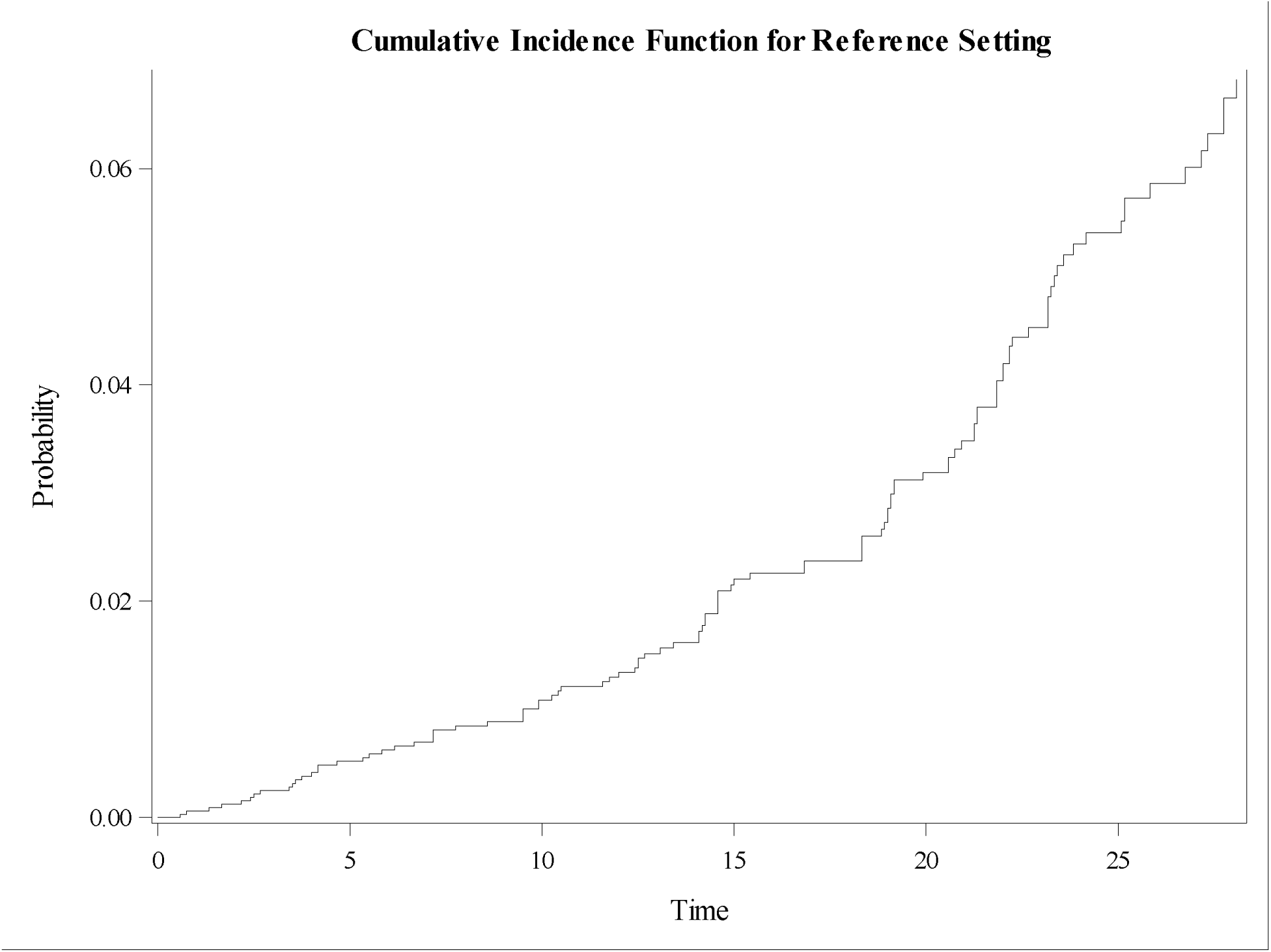
Cumulative Incidence Function (CIF) of Periodontal Loss over a 30 Year Follow Up Period

In addition, we fitted four commonly used parametric models, namely, Weibull, Exponential, Log-logistic, and Log-normal to the baseline hazard function λ_0_ (t) and computed hazard rates for the covariates based on the CPH model. Table 7 provides the results based on these four models.

**Table 7:** Hazard Rate based on Parametric Survival Regression Models.

|  | Weibull |  |  | Exponential |  |  |
| --- | --- | --- | --- | --- | --- | --- |
| Characteristics | Estimate of Beta | 95% CI | p-value | Estimate of Beta | 95% CI | p-value |
| Age | 0.0006 | -0.003,0.005 | 0.7726 | 0.0030 | -0.008,0.014 | 0.5880 |
| Gender | 0.0943 | -0.012,0.200 | 0.0812 | 0.2030 | -0.090,0.496 | 0.1744 |
| Tooth Type |  |  |  |  |  |  |
| Non-Molar | - | - | - | - | - | - |
| Upper Molar | 0.1511 | 0.002,0.300 | 0.0470 | 0.3915 | -0.023,0.807 | 0.0644 |
| Lower Molar | 0.1027 | -0.040,0.245 | 0.1583 | 0.1314 | -0.256,0.519 | 0.5060 |
| Maintenance | 0.0431 | 0.039,0.047 | <0.0001 | 0.0779 | 0.070,0.086 | <0.0001 |
| MM Class |  |  |  |  |  |  |
| 2 | - | - | - | - | - | - |
| 3 | 0.2466 | 0.117,0.377 | 0.0002 | 0.4502 | 0.101,0.800 | 0.0116 |
|  | Log-logistic |  |  | Log-normal |  |  |
| Age | 0.0011 | -0.003,0.006 | 0.6351 | 0.0008 | -0.005,0.006 | 0.7660 |
| Gender | 0.1293 | 0.011,0.248 | 0.0327 | 0.1691 | 0.030,0.308 | 0.0173 |
| Tooth Type |  |  |  |  |  |  |
| Non-Molar | - | - | - | - | - | - |
| Upper Molar | 0.1039 | -0.056,0.264 | 0.2035 | 0.1231 | -0.069,0.315 | 0.2083 |
| Lower Molar | 0.0599 | -0.095,0.215 | 0.4483 | 0.0209 | -0.165,0.207 | 0.8256 |
| Maintenance | 0.0448 | 0.041,0.049 | <0.0001 | 0.0490 | 0.045,0.053 | <0.0001 |
| MM Class |  |  |  |  |  |  |
| 2 | - | - | - | - | - | - |
| 3 | 0.3578 | 0.213,0.502 | <0.0001 | 0.3741 | 0.207,0.542 | <0.0001 |

**Table 8:** Hazard Rates based on Cox and Parametric Survival Regression Models.

|  | Cox Proportional<br>Hazard | Weibull | Exponential | Log-logistic | Log-normal |
| --- | --- | --- | --- | --- | --- |
| Age | 0.99 | 1.00 | 1.00 | 1.00 | 1.00 |
| Gender | 1.12 | 1.10 | 0.82 | 0.88 | 0.84 |
| Teeth Upper Molar | 1.13 | 1.16 | 0.68 | 0.90 | 0.88 |
| Teeth Lower Molar | 1.48 | 0.90 | 0.88 | 0.94 | 0.98 |
| Maintenance | 0.88 | 1.04 | 0.92 | 0.96 | 0.95 |
| Miller Group 3 | 2.37 | 0.78 | 0.64 | 0.70 | 0.69 |
| AIC |  | 615.4 | 810.4 | 618.2 | 653.3 |

### Parametric Survival Models

Competing risk models provide more precise probability of death when competing events are present, as opposed to the standard survival models. Frequently pointed out that, in presence of competing events, standard product limit (KM) method of estimating survivor function or the PH model yield biased results for event of interest as the probability of death or loss of tooth is modified by an antecedent competing event. Among the four commonly used parametric models in medical research (Weibull, Exponential, Loglogistic, and Log-normal), the Weibull model had the lowest value of Akaike Information Criterion (AIC) to be 615.4. Low AIC value is an indicator of a good fit of the model to the data.

Note that based on the selected Weibull model, both the prognosticator maintenance and MM class III are significant but the estimated hazard for MM class III is 0.78.

Based on Tables S1 and S2, the risk of losing a tooth decrease over time irrespective of MM class. One interesting observation, in our dataset, is that within the first five years, most of the periodontal loss happened within the first five years as compared to other loses (69.2% vs. 30.8% for MM class II) and (77.8% vs. 22.2% for MM class III. See supplement.

## Discussion

The use of survival analysis is limited in dental research, especially the application of competing risk survival models. For cost-effective/cost-utility studies, primarily, it is necessary to estimate the precise probability of an event, i.e., losing a tooth, either at a specific time (t) or within a pre-specified time interval. This estimate of probability is affected by other causes of failure other than the cause of failure of interest. Accounting for other causes of failure while estimating the probability of failure of an event of interest is handled through competing risk models. In this current work, we have demonstrated the hazard rate estimate based on a basic Kaplan-Meier survival method. This is a non-parametric method of estimating survival or mortality curves without including the effects of prognosticators. Next, we introduced covariates like age, gender, teeth type, number of maintenance visits, and severity of the tooth condition via Miller-McEntire classification with the help of the CPH model. We estimated the hazard rate or probability of losing a tooth with periodontal disease over time by the CPH cause-specific (CS) model and then by the subdistribution FG model. In the cause-specific model, we treated non-periodontal loss as a censored event, which resulted in an overestimation of the CIF but in the sub-distribution model, we treated non-periodontal loss as part of the risk set and found an estimate of the hazard rates to be lower than the one from the CS model. This means our overestimation of the benefits of periodontal therapy by the FG model would in turn imply the relatively higher cost-effectiveness of supportive periodontal therapy (SPT). Overestimation of hazard by cause-specific competing risk would have profound consequences for any cost-effectiveness/cost-utility study. To the best of our knowledge, this is the first study where a precise estimate of probability of losing a tooth due to periodontitis is suggested. Also, the issue of how close the number of competing event (s) are to the number of events of interest would affect the hazard rate, and so would the cost estimates. If the number of competing events is much closer or exceeds the number of events of interest, then it would limit the clinical usefulness of the costly intervention to treat the disease of interest, in our case, periodontal disease. Initially, if the number of competing events is much closer to the number of events of interest, then the estimated hazard rate of the event of interest will be higher than otherwise. In our research, we have not explored this aspect, but it is an open area of research. We also computed hazard rate estimates based on parametric models and found comparable results to that of semi parametric models except for the MM class. In our work, we found comparable results but both the approaches, one based on semi parametric model and one on criterion-based parametric model. It is the baseline hazard λ_0_ (t) that is modeled using parametric models like exponential, Weibull, log-logistic, and log-normal. It is quite possible that some other parametric model may be a better fit than these four commonly used parametric model and may provide hazard rate estimate for MM class III to be like that obtained by the subdistribution model. One would prefer to use parametric models only if any of the chosen parametric models fit the data well. The issue of fitting parametric models to baseline hazard is complex and may require fitting a separate parametric model for the periodontal loss and a different parametric model for the non-periodontal loss. This is a research area which can be explored in future work.

We did not discuss the effect of time dependent covariates. Suppose a blood marker for periodontal disease which varies over time can be incorporated in the model. Such a model can be easily implemented in the framework of CS and SD models using current software. We also did not explore the stratum effect i.e., age can be divided into different stratum and one can fit these models by stratum. We encourage researchers to think about these issues for their datasets so ultimately the cost of periodontal care over time can be more precisely estimated. Cost estimates utilizing the probabilities of a periodontal loss under different scenarios will be part of our future work.

## Conclusion

Competing risk models should be used whenever there are two or more events under study. Cumulative Incidence Function provides the correct estimates of cumulative hazard over time when competing events are at play. More precision to the estimate of hazard function can be added by using parametric distributions to the baseline hazard rate provided one of the parametric models fits the data well. This study advocates for the application of competing risk models and emphasizes the accuracy of the Cumulative Incidence Function in estimating cumulative hazard over time, particularly in the context of tooth loss due to periodontitis and the associated long-term dental care costs. The use of parametric distributions further enhances precision in estimating the hazard function, providing a foundation for more nuanced risk assessments over extended follow-up periods.

## Data Availability

All data produced in the present study are available upon reasonable request to the authors.

## Declarations

Availability of data and materials

The dataset used and analyzed during the current study is available from the corresponding author on reasonable request.

## Author’s Contribution

HD and MS conceived the project and provided valuable input in all the clinical aspects of the study and in writing of the manuscript. AS has provided valuable input, conducting the study, analysis, writing the initial draft, reviewing, and editing the article. SNR has contributed towards reviewing the article with constructive input in writing and analysis of data. AB and PS have contributed towards data analysis and aided in drafting the paper.

## Acknowledgments

The authors would like to acknowledge the Department of Diagnosis and Oral Health, University of Louisville School of Dentistry, KY, USA for the funding to conduct the research, and the reviewers for their insightful comments and time.

## Supplement

**Table S1:** Number of Teeth Loss during SPT (MM Class II) by Years of Follow UP.

| Status of Tooth | < 5 | 5-10 | 10-15 | 15-20 | 20-25 | 25-30 | ≥30 | Total |
| --- | --- | --- | --- | --- | --- | --- | --- | --- |
| Survived | 0 | 0 | 16<br>(32.7) | 84<br>(79.3) | 176<br>(80.4) | 184<br>(92.9) | 180<br>(93.8) | 640 |
| Perio Loss | 9<br>(69.2) | 11<br>(57.9) | 16<br>(32.7) | 10<br>(9.4) | 22<br>(10.1) | 5<br>(2.5) | 6<br>(3.1) | 79 |
| Other Loss | 4<br>(30.8) | 8<br>(42.1) | 17<br>(34.7) | 12<br>(11.3) | 21<br>(9.6) | 9<br>(4.6) | 6<br>(3.1) | 77 |
| Total | 13 | 19 | 49 | 106 | 219 | 198 | 192 | 796 |

**Table S2:**
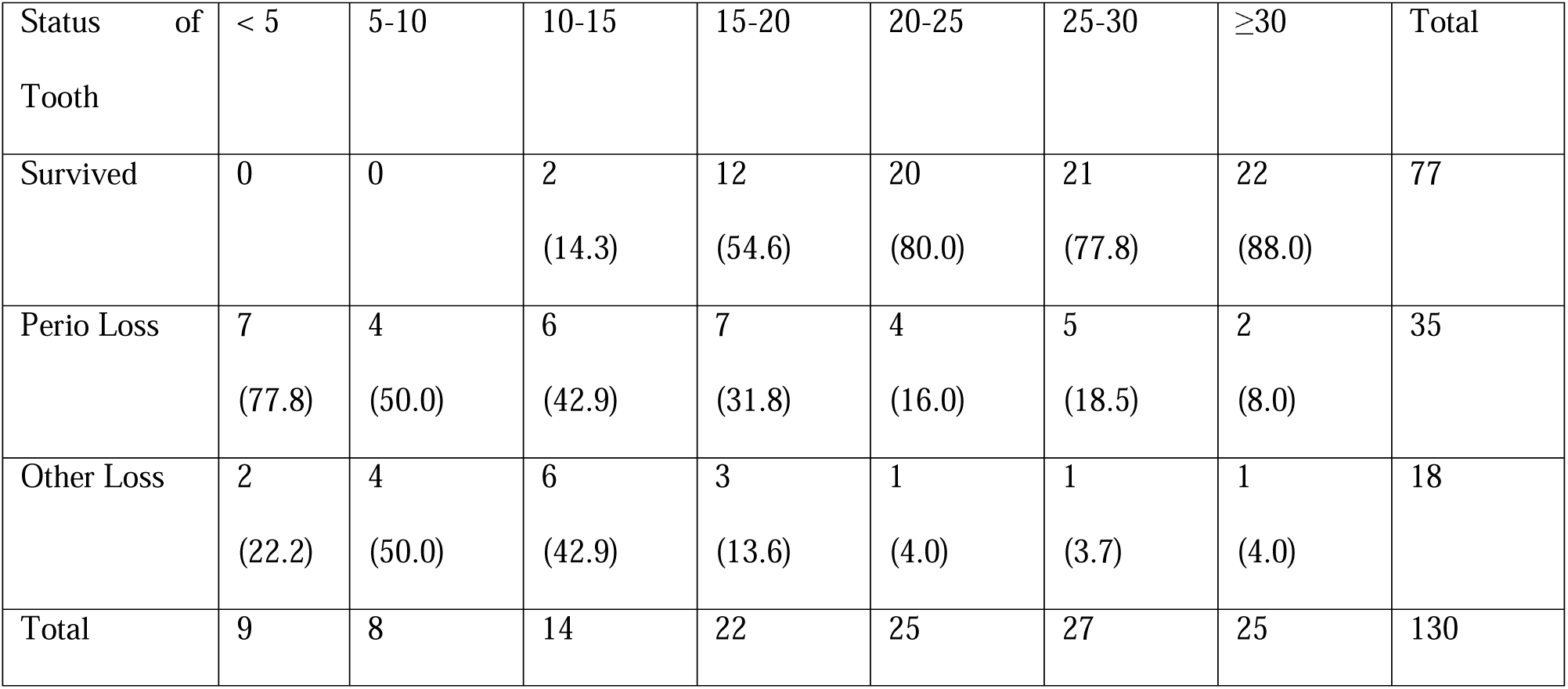
Number of Teeth Loss during SPT (MM Class III) by Years of Follow UP.

| Status of Tooth | < 5 | 5-10 | 10-15 | 15-20 | 20-25 | 25-30 | ≥30 | Total |
| --- | --- | --- | --- | --- | --- | --- | --- | --- |
| Survived | 0 | 0 | 2<br>(14.3) | 12<br>(54.6) | 20<br>(80.0) | 21<br>(77.8) | 22<br>(88.0) | 77 |
| Perio Loss | 7<br>(77.8) | 4<br>(50.0) | 6<br>(42.9) | 7<br>(31.8) | 4<br>(16.0) | 5<br>(18.5) | 2<br>(8.0) | 35 |
| Other Loss | 2<br>(22.2) | 4<br>(50.0) | 6<br>(42.9) | 3<br>(13.6) | 1<br>(4.0) | 1<br>(3.7) | 1<br>(4.0) | 18 |
| Total | 9 | 8 | 14 | 22 | 25 | 27 | 25 | 130 |

